# The pitfalls of incidence-based time series regression for inferring the effects of weather on infectious diseases

**DOI:** 10.64898/2026.03.13.26348326

**Authors:** Pietro Gemo, Laura Andrea Barrero Guevara, Cana Kussmaul, Sarah C. Kramer, Matthieu Domenech de Cellès

## Abstract

A central question in environmental epidemiology is how the weather affects infectious diseases. Time-series regression (TSR) on population-level case incidence data is widely used to estimate weather effects; however, this design may be biased due to the complexities of infectious disease dynamics, including nonlinear feedback, various types of noise, and latent, dynamic variables such as population immunity. Here, we assess the reliability of incidence-based TSR through a controlled simulation study across four different climates and fifty scenarios representing different pathogens. For each scenario, we simulated 10 years of weekly incidence data using a simple transmission model that included real-world weather data on temperature and relative humidity. We then examined whether the ground-truth weather effects could be recovered from model simulations using negative binomial generalized additive models, a flexible class of TSR models commonly used in empirical applications. We find that these models frequently fail to yield accurate and precise estimates of weather effects, even under favorable conditions such as no process noise and low observation noise (overdispersion). Hence, our results caution against the indiscriminate use of TSR models and suggest that more mechanistic approaches are needed for statistical inference of weather effects from population data.

## 2 Introduction

A key research question in environmental epidemiology is how weather—and by extension climate— affects infectious diseases [1]. Mechanistically, weather effects are supported by experimental evidence showing that variables such as temperature and relative humidity can sensitively alter the survival time, infectivity, or virulence of many pathogens. These include, among others, respiratory viruses such as SARS-CoV-2 [2] or influenza [3], bacteria such as *Streptococcus pneumoniae* [4] or *Vibrio cholerae* [5], and malaria parasites [6]. Beyond its effects on the pathogen, weather can also impact transmission and infection through other mechanisms operating at different scales, from within-host (e.g., low relative humidity impairing mucociliary clearance of influenza A viruses in mice [7]) to between-host effects (e.g., temperature-related seasonal variations in human contact patterns [8]).

Although such evidence provides a solid foundation for postulating the effects of weather on infectious diseases, observational research remains necessary to quantify these effects at the population level. However, previous research has identified several methodological challenges in inferring these effects from case incidence data [1,9,10]. First, given the observational nature of these data, careful analysis is necessary to distinguish correlation from causation. Second, surveillance typically detects only a limited number of cases, leading to measurement noise and incomplete data. Third, the infection process underlying the data is latent and dynamic; key variables, such as the size of the immune population, are unobserved but may fluctuate rapidly and strongly influence transmission dynamics [1]. Altogether, these challenges may complicate the use of standard statistical methods that ignore the disease’s biology.

The time-series regression (TSR) study design may be vulnerable to these issues. In this design, the time series of incident cases is regressed against weather variables, with the resulting regression coefficients interpreted as causal effects. Often, additional covariates are included to capture intrinsic drivers of infection, e.g., autocorrelation terms to model transmission effects or smooth functions of time to model changes in the immune population [9]. Due to its simplicity and intuitive appeal, the TSR design is widely used in environmental epidemiology [10,11], yet its reliability remains unclear because of the aforementioned challenges.

In our previous work, we argued that TSR estimates of the weather effects may be subject to measurement bias [12]. As shown in Fig. 1, this bias arises from systematic differences between the unobserved target endpoint, directly affected by weather variables (i.e., the transmission rate), and the observed endpoint (i.e., the incidence rate). Theoretically, additional covariates could account for unobserved variables along the causal path linking the two endpoints, thereby mitigating this bias. Still, it remains unclear whether such additions work well in practice. Here, we build on this prior work to systematically evaluate the performance of TSR studies. Using an extensive simulation study based on a simple transmission model, we aimed to evaluate the accuracy and precision of TSR estimates compared to a fixed ground truth. We repeated this procedure to quantify changes in TSR performance across varying experimental treatments [13], including climate, pathogen characteristics, observation and process noise levels, and extrinsic seasonal drivers.

**Figure 1:**
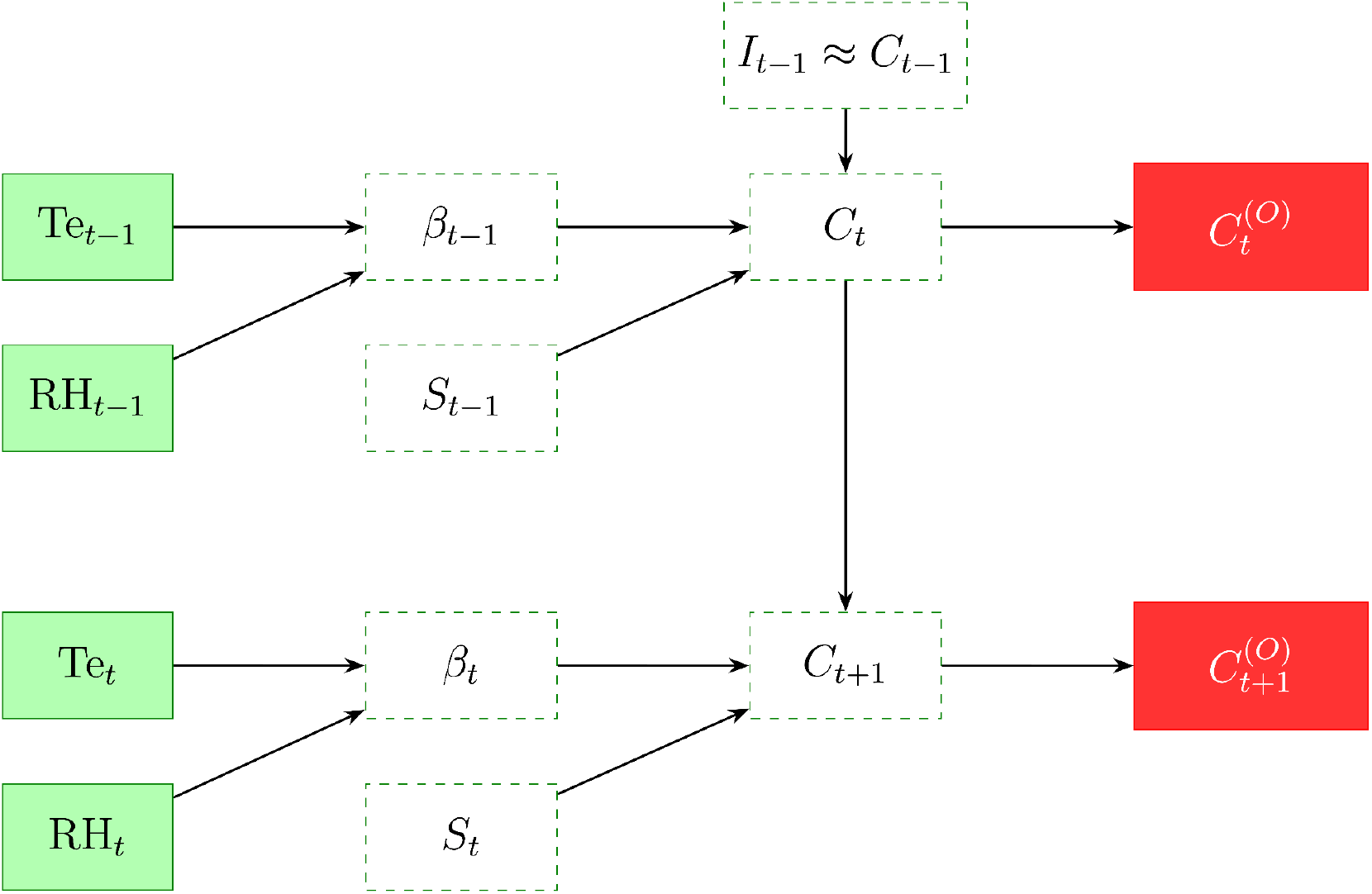
Causal directed acyclic graph for the transmission model. Green boxes represent the observable climatic variables. Dashed boxes show the unobserved states of the system. *Te*_*t*_ and *RH*_*t*_ are temperature and relative humidity respectively, *S*_*t*_ represents the number of susceptible individuals, *β*_*t*_ is the transmission rate, 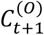 is the weekly number of new observed cases, and *C*_*t*_ is the total number of cases. Due to the chosen transmission model, the latter variable is approximately equal to the total number of infected *I*_*t*_.

## 3 Methods

### 3.1 Overview & Modeling workflow

To assess TSR’s performance, we conducted a simulation study. Our modeling workflow is illustrated in Fig. 2. First, we designed a simple epidemic model that incorporated real-world weather data, which were assumed to modulate the transmission rate seasonally (see Fig. 1 for the directed acyclic graph (DAG) underlying the epidemic model). We then used this model to simulate time series of weekly reported cases, an outcome comparable to that used in TSR studies. Finally, we fitted different TSR models to these time series and assessed their ability to capture the real effects of weather variables on transmission. For comprehensiveness, we repeated this process for weather data across four different climates and under various assumptions about pathogens’ transmissibility and the rate of waning of infection-derived immunity. The steps of the modeling workflow are detailed below.

**Figure 2:**
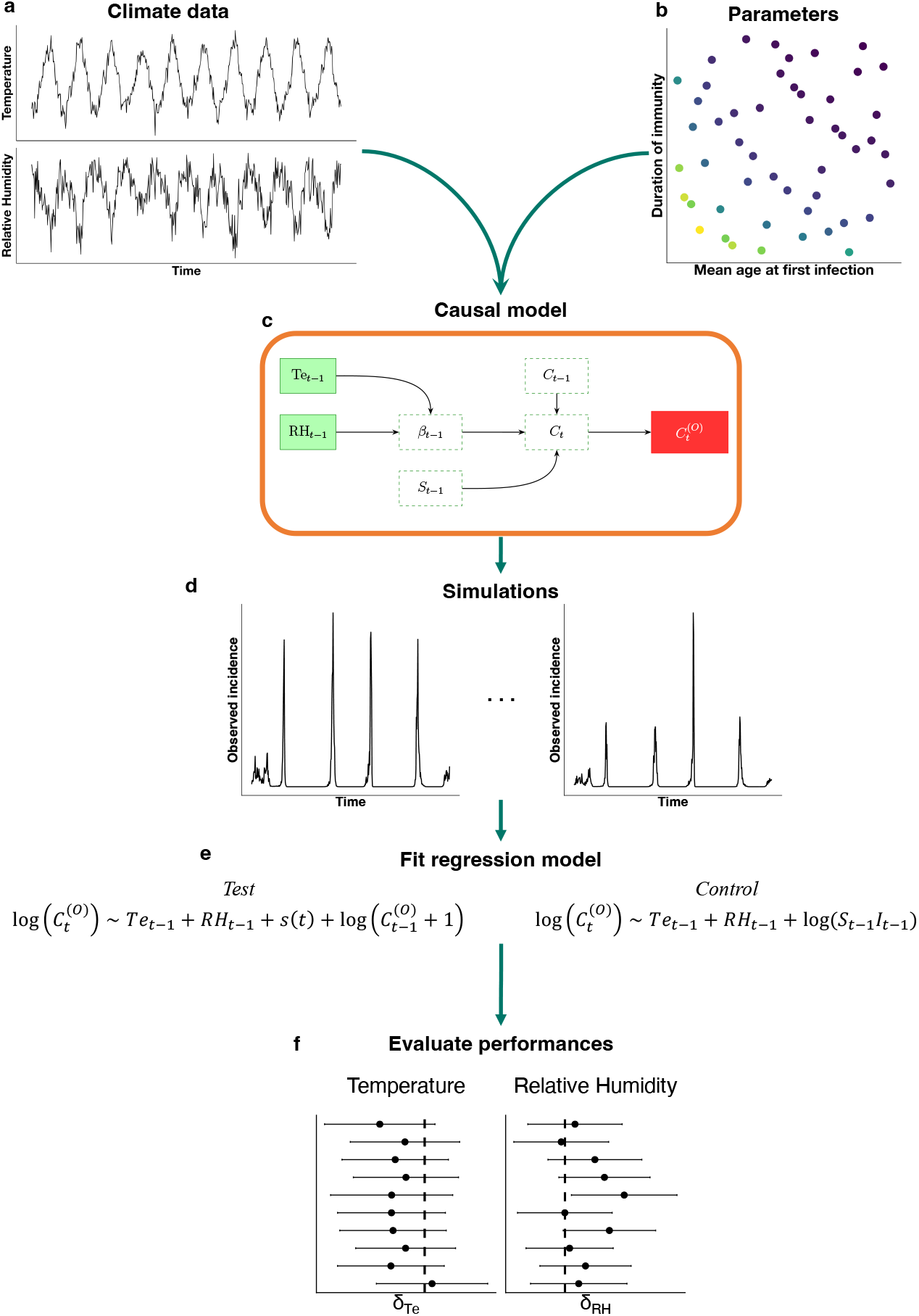
Workflow of the simulation study. a) Time series of temperature and relative humidity measurements. b) Set of sampled values of the mean age at first infection and immunity waning rate fed to the causal model. c) DAG of the causal model, here defined as a simple transmission (SIRS) model. d) 100 replicate time series are generated for each set of sampled parameters. e) The two (control and test) GAM models are fitted to each simulated time series. f) The accuracy and precision of estimates are evaluated in comparison to the ground-truth values of parameters.

### 3.2 Climatic data

#### 3.2.1 Location selection

To ensure sufficient global coverage of climates, we selected four locations (Supplementary Table S1) representing the main categories of the Köppen-Geiger climate classification system [14]:

1. Rio de Janeiro, Brazil, which exhibits a tropical savanna climate characterized by warm to hot temperatures throughout the year and distinct wet and dry seasons (Aw Köppen-Geiger code);
2. Dubai, United Arab Emirates, classified as a hot desert climate, marked by hot, arid conditions with intense sunshine and minimal annual precipitation (BWh);
3. Rome, Italy, a hot-summer Mediterranean climate, with hot, dry summers and mildly wet winters (Csa);
4. Toronto, Canada, whose continental climate is associated with warm summers, cold winters, significant seasonal temperature variations, and largely constant relative humidity (Dfb).

#### 3.2.2 Data source

The weather data were available from the “Prediction Of Worldwide Energy Resource” project, initiated by NASA, and extracted using the R “nasapower” package (version 4.2.2)[15]. The data consisted of daily satellite-based temperature and relative humidity measurements from 2010 to 2020. We averaged these data weekly to yield 520 weekly averages, which we incorporated into our transmission model (Supplementary Fig. S1).

### 3.3 Model formulation

#### 3.3.1 Transmission rate & Weather effects

To incorporate weather effects into our model, we assumed that temperature (*Te*_*t*_) and relative humidity (*RH*_*t*_) each directly affected the transmission rate (*β*_*t*_). This assumption is supported by experimental evidence showing that both weather variables affect the survival of respiratory viruses [3,10,16–19]. Specifically, the transmission rate was modeled by the following equation:

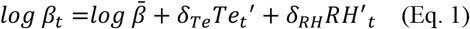

Here, 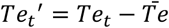 represents the mean-centered temperature at time *t* and 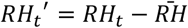 the mean-centered relative humidity at time *t*, where 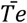 and 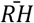 are the temporal averages across the entire time series. The parameter 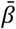 represents the mean transmission rate, defined as 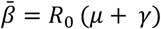, where *R*_0_ is the basic reproduction number, *μ* is the birth and death rate, and *γ* the generation rate (for a complete description of all model parameters, refer to Supplementary Table S2). The parameters *δ*_*Te*_ and *δ*_*RH*_ capture the direct causal effects of temperature and relative humidity on transmission rate.

#### 3.3.2 Transmission model

Following Ref. [12], we formulated a discrete-time SIRS model with demography. We assumed a fixed time step and a generation time of one week, thereby enabling us to specify a control TSR model that recovered the true causal effects of weather when all model variables were perfectly observed (see the “Control” model section below). Given these assumptions, the model was represented by the following set of difference equations:

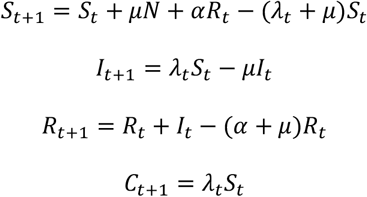

where the parameter *N* is the population size, *μ* the birth/death rate (assumed equal to maintain a constant population over time), and *α*^−1^ the average duration of infection-derived immunity. The force of infection is defined as *λ*_*t*_ = *β*_*t*_(*I*_*t*_ + *ι*)/*N*, where *ι* = 10^−5^ denotes the inflow of a small number of infected individuals introduced to prevent epidemic extinction. Finally, *C*_*t*_ is the total number of new cases every week (total incidence rate). For added biological realism, we introduced stochasticity in the transmission rate: at each time step, we multiplied *β*_*t*_ by a random deviate *δW*_*t*_ from a Gamma distribution (*Γ*_*WN*_) with mean 1 and standard deviation *σ*_*β*_. Hence, the parameter *σ*_*β*_ modulates the magnitude of stochasticity in transmission, and in the limit *σ*_*β*_ → 0 the transmission model is entirely deterministic. Altogether, the force of infection was modeled as:

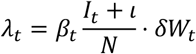

#### 3.3.3 Observation model

To align with the outcomes analyzed in TSR studies, we defined the weekly time series of new observed cases (i.e., the observed incidence rate, denoted by 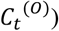 as the primary outcome of the model. We generated this outcome using a Negative-Binomial observation model:

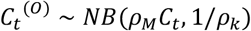

with mean *μ*_*t*_ = *ρ*_*M*_*C*_*t*_ and variance *μ*_*t*_ + *ρ*_*K*_*μ*_*t*_^2^. Here, the parameter *ρ*_*M*_ is defined as the reporting probability and *ρ*_*K*_ the reporting overdispersion. Hence, this observation model enabled us to generate measurement noise in the simulated data, as expected in real-world data settings. In the baseline parameterization, we set *ρ*_*K*_ to 0.1, corresponding to a small coefficient of variation for the observation model. We note that this value is lower than the minimum estimated by transmission models across a range of pathogens (see illustrative values from a non-exhaustive search in Supplementary Table S3).

### 3.4 Model parameterization

#### 3.4.1 Parameterization of pathogens’ characteristics

To make our analysis more generalizable, we considered a range of pathogens with different average durations of immunity (parameter *α*^−1^, range considered: 1–20 years) and mean age at first infection (parameter *A*, range considered: 1–10 years). For our model, the latter parameter is defined as:

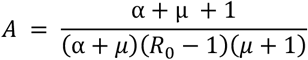

Specifically, we used Latin hypercube sampling (over the parameter ranges indicated above) to define 50 synthetic pathogens with distinct pairs of (*α*^−1^, *A*) parameters (Supplementary Fig. S2). We chose these specific parameters for their clear biological interpretation.

#### 3.4.2 Calibration of weather effects on transmission

To derive realistic values for the weather effect parameters (*δ*_*Te*_ and *δ*_*RH*_), we calibrated them to reproduce the seasonal variations in transmission rates estimated in previous modeling studies. Specifically, we considered two modeling studies: one that estimated a ±15% seasonal amplitude (relative to the mean) in influenza transmission in Tel Aviv, Israel [20], and another that estimated a

±37.5% seasonal amplitude in RSV transmission in Turku, Finland [21]. Using the observed weather data from both locations during the corresponding study periods and assuming that *δ*_*Te*_ = *δ*_*RH*_ = *δ*, we estimated the weather parameter that yielded the best match in relative amplitude between the simulated and empirical transmission rates. The resulting estimates were *δ* = −0.02 in Tel Aviv and *δ* = −0.04 in Turku, and we considered both values in subsequent analyses.

#### 3.4.3 Model versions and alternative model formulations

We defined the baseline model version as the model with no process noise (*σ*_*β*_ = 0), low observation noise (*ρ*_*K*_ = 0.1), and the strongest weather effects (*δ* = −0.04). We tested four other model versions by varying different parameters:

1. Increased observation noise (*ρ*_*K*_ = 0.16)
2. Addition of process noise (*σ*_*β*_ = 0.05)
3. Decreased weather effects (*δ* = −0.02)
4. Addition of term-time forcing in transmission.

For the latter version, we formulated an extended model with an additional source of seasonality in transmission: seasonal variations in host contacts, driven by the alternation of school terms (with higher contact rates) and school holidays (with lower contact rates). This effect was modeled as a multiplicative change *School*(*t*) in the transmission rate, proportional to (1 − *δ*_*S*_) during school holidays (defined here as Christmas and summer vacations, weeks 52 and 24–35) and (1 + *δ*_*S*_) during school terms. Here, *δ*_*S*_ represents the amplitude of term-time forcing in transmission, fixed under the assumption that children make 30% fewer contacts during school holidays [22–25]. Accordingly, for time series including term-time forcing, the regression models included a holiday indicator as a covariate, indicating whether the observation fell during a school holiday.

### 3.5 Regression models formulation and fitting

The TSR models were formulated as Generalized Additive Models (GAMs) with a Negative Binomial distribution to match the observation model. Although GAMs are not universally used in TSR studies, we considered this class of regression models for their flexibility and versatility, particularly for capturing continuous temporal variation while preventing overfitting. We note that many TSR models used in the literature (e.g., [26–31]) are just special cases of GAMs. For each simulated time series of observed incidence, we formulated and fitted two GAMs, as described below.

#### 3.5.1 Control model

Following Ref.[12], we first identified a suitable TSR model to apply to the outputs of our transmission model. To do so, we log-transformed the total incidence rate:

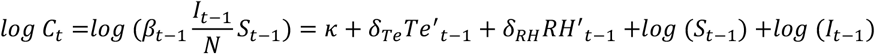

where *κ* is a constant. This equation, alongside the NB observation model for the observed incidence rate, indicates that an exact TSR model in our setting is an NB model with log-link and three covariates: *Te*^′^_*t*−1_, *RH*^′^_*t*−1_, and *log* (*S*_*t*−1_) +*og* (*I*_*t*−1_). Hereafter, we refer to this model as the “Control” model, as we expect it to recover the true causal effects of weather. We note that this model is conceptually equivalent to the Time-series Susceptible-Infected-Recovered (TSIR) model, a regression-based approach where the reconstruction of the susceptible population is informed by a transmission model [32].

#### 3.5.2 Test model

In real-world settings, the variables *log S*_*t*−1_ and *log I*_*t*−1_ are unobserved and must be approximated by adding covariates to the TSR model. Hence, we considered a TSR test model with two additional covariates. The first additional covariate was an autoregressive term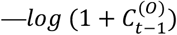. This addition was based on the DAG (Fig. 1) and the fact that, in our simple model, *I*_*t*_ ≈ *C*_*t*_. Hence, the autocorrelation term may approximately capture temporal variations in the number of infected. More broadly, including an autoregressive term is generally recommended in TSR models to account for the autocorrelation in the outcome time series [10].

The second additional covariate is a smooth function of time, modeled using a thin plate regression spline. This choice follows the recommendations for the TSR model design [9,10,33] and reflects the practice of multiple TSR studies [29–31,34–40]. We set the maximum basis dimension to 52 (approximately one degree of freedom per 10 data points) to provide this covariate with sufficient flexibility to capture temporal variation in the unobserved variable *log* (*S*_*t*−1_). This choice was based on preliminary analyses showing that the effective degrees of freedom estimated for this smooth tended to saturate at this value. However, we tested a higher value of 75 in a sensitivity analysis.

#### 3.5.5 TSR performance assessment

To assess each TSR model’s ability to capture the causal effects of weather, we computed two average performance metrics for each combination of weather variable, scenario, and location:

1. **Relative Mean Absolute Bias (RMAB)**. We first calculated the Mean Absolute Bias as the absolute difference between the estimated and true weather effects, averaged across within-scenario replicates. The RMAB was then defined as the scenario-specific MAB divided by the absolute value of the true weather effect. For interpretation, an RMAB of 20% means that estimates were on average 20% off the actual weather effect value. Hence, the RMAB quantifies the average estimation accuracy.
2. **Relative Mean Standard Error (RMSE)**. This metric was calculated to allow comparison in precision across tested model versions. For each scenario, we first averaged the standard errors of the estimates across replicates to obtain the Mean Standard Error (MSE). Then, we divided each MSE by the absolute value of the true weather effect. For interpretation, an RMSE of 10% means that the average estimation uncertainty was ±10% relative to the true weather value effect.

In addition, we evaluated each metric’s standard deviation across scenarios (denoted by *σ*_*I*_) and replicates (denoted by *σ*_*J*_) within each scenario; see supplementary material for the corresponding equations.

Finally, we also calculated the mean power for every scenario. This was defined as the average across replicates of a binary variable equal to 1 if the sign of the weather effect was correctly inferred (i.e., if the entire 95% confidence interval contained negative values) and 0 otherwise. Hence, this less stringent metric assessed the TSR models’ ability to correctly capture the sign of the weather effect.

### 3.6 Simulation protocol

In every city and for every parameter pair (representing the pathogen’s characteristics), we simulated 100 stochastic replicates (Fig. 3). We then recorded the simulated time series of the observed incidence rate and fitted the control and test TSR models using these time series as the outcome. All the models were initialized at the steady state of the transmission model without seasonal forcing.

**Figure 3:**
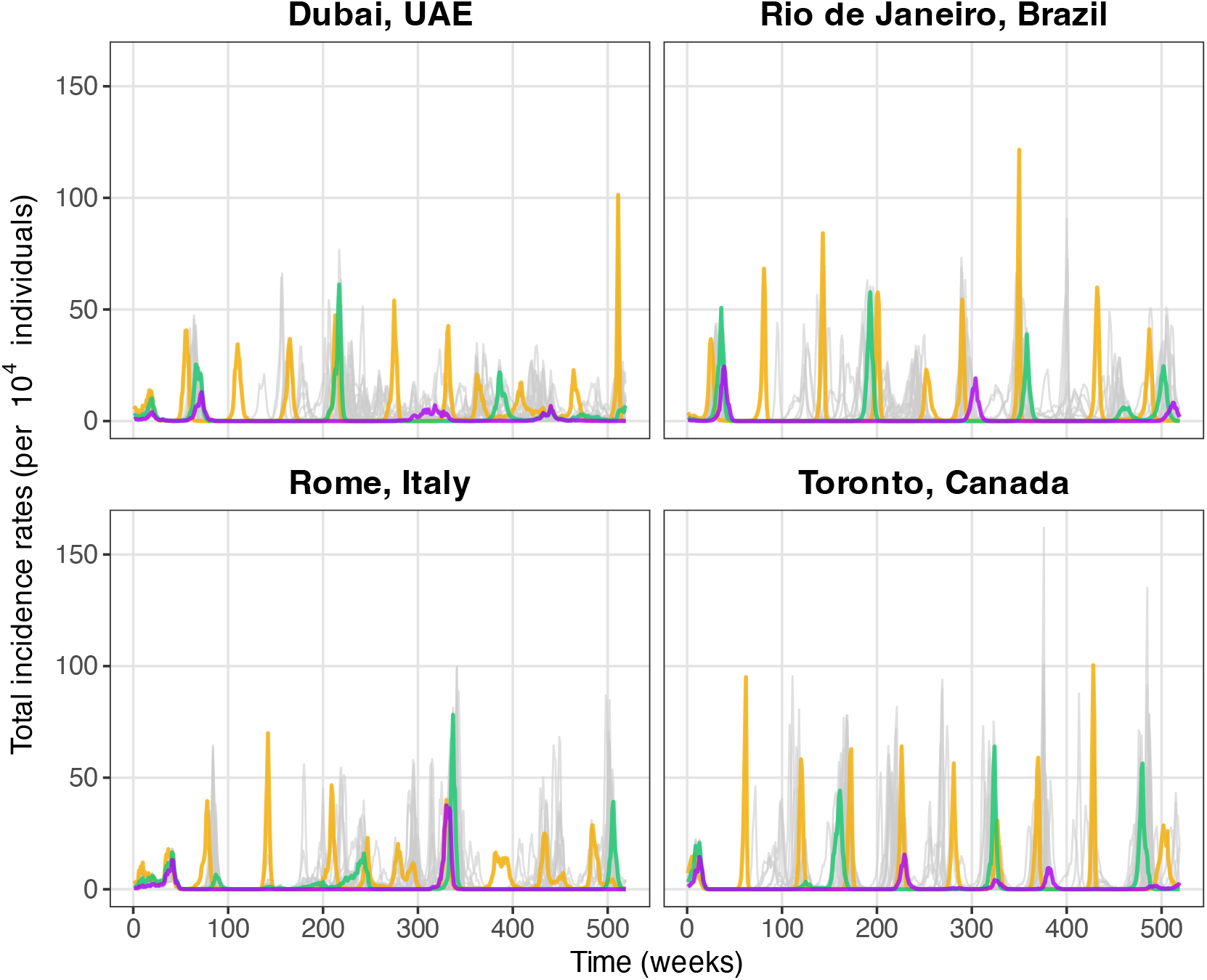
Simulated time series of the total incidence rate. The colored lines correspond to three selected scenarios, highlighted to illustrate the different epidemic periodicities, while all remaining simulations are shown in grey. These simulations refer to the baseline scenario (*δ*_*Te*_ = *δ*_*RH*_ = −0.04, *ρ*_*M*_ = 50*%, ρ*_*K*_ = 10*%, σ*_*β*_ = 0*%*).

### 3.7 Numerical implementation

All the transmission models were formulated and simulated using the R package *pomp* (version 6.1) [41]. The TSR models were fitted using the R package *mgcv* (version 1.9.1) [33], and all analyses were implemented in R version 4.4.3.

## 4 Results

We first verified that the control model produced accurate and precise estimates of weather effects, with both RMAB and RMSE around 10% at all locations except Rio de Janeiro, with an average RMAB of 17% and average RMSE of 21% across weather variables (Supplementary Table S4, Fig. 4–5). In addition, all control models had 100% power to detect the correct sign of both weather variables.

**Figure 4:**
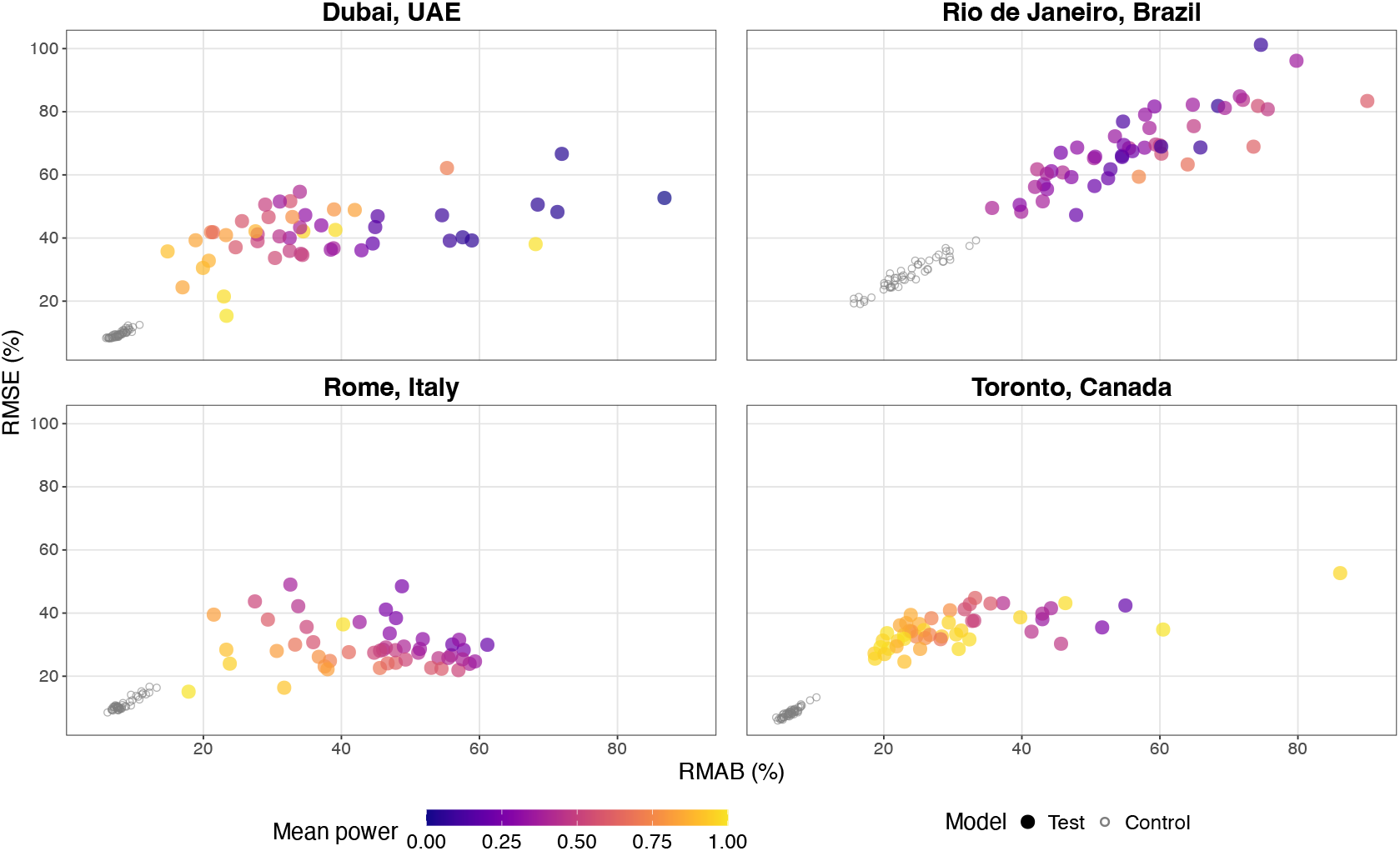
Assessment of TSR models’ performance in estimating the effect of temperature. Each point represents a tested synthetic pathogen, with TSR performance quantified by the Relative Mean Absolute Bias (RMAB; x-axis, averaged across replicates) and the Relative Mean Standard Error (RMSE; y-axis, averaged across replicates). The color scale represents the average power, defined as the probability that the estimated parameter correctly recovers the sign of the true effect. The grey points indicate the control model’s performance. The parameters used to generate the fitted simulations were: *δ*_*Te*_ = *δ*_*RH*_ = −0.04, *ρ*_*M*_ = 50*%, σ*_*β*_ = 0*%, ρ*_*K*_ = 10*%*.

The estimation results for the baseline test model formulation are presented in Figs. 4 and 5. Overall, the temperature effect estimates were inaccurate across all locations, with the mean RMAB ranging from 32% in Toronto to 57% in Rio de Janeiro. Furthermore, there was substantial RMAB variability across scenarios (*σ*_*I*_ range across locations: 11–16%) and across stochastic replicates within scenarios (*σ*_*J*_ range across locations: 22–42%). In addition, there was marked uncertainty in the estimates, with mean RMSE ranging from 29% in Rome to 68% in Rio de Janeiro. The RMSE varied among (*σ*_*I*_ range: 6–12%) and within (*σ*_*J*_ range: 2–5%) scenarios. Finally, average statistical power was low, ranging from 78% in Toronto to 35% in Rio de Janeiro.

**Figure 5:**
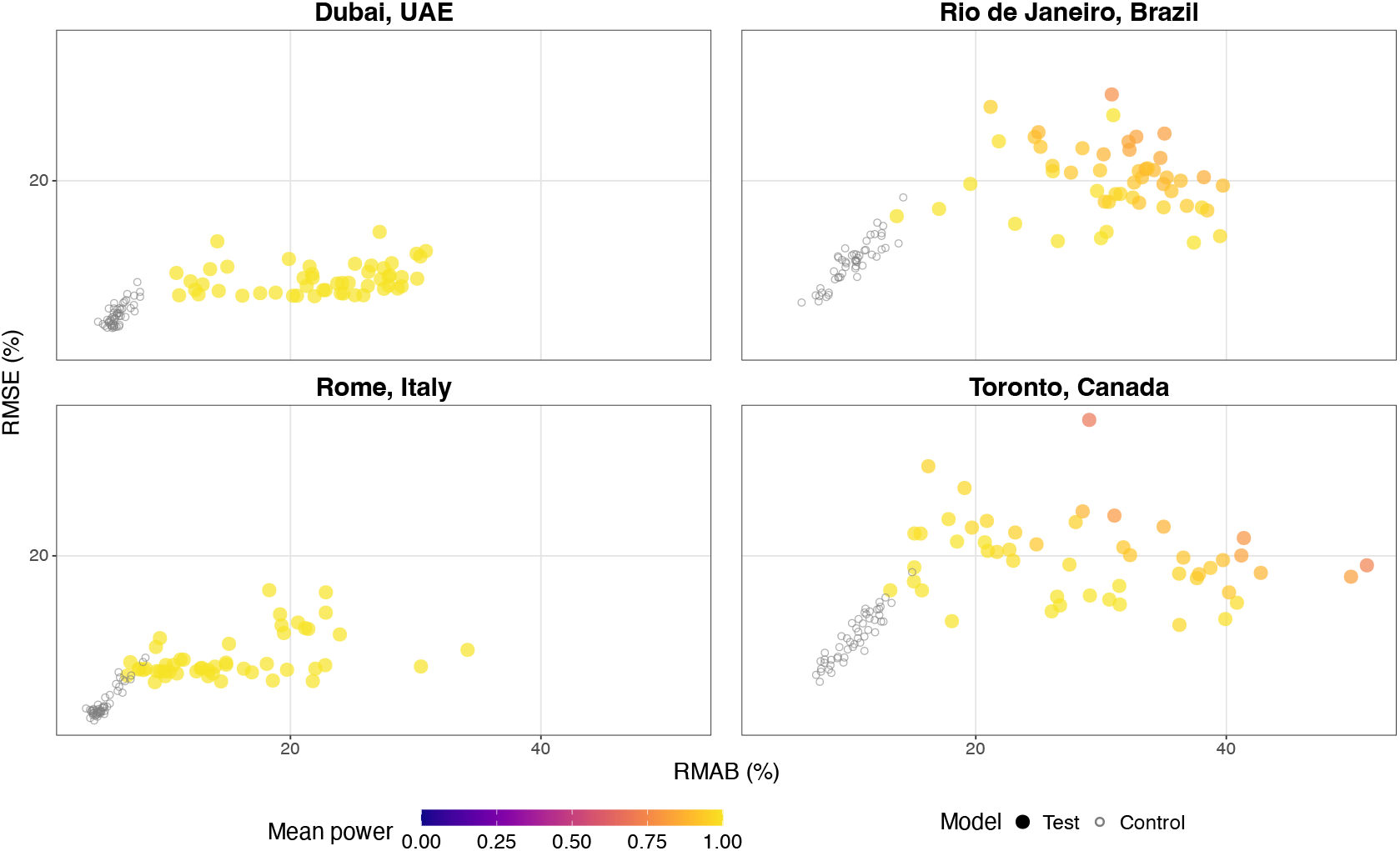
Assessment of TSR models’ performance in estimating the effect of relative humidity. Each point represents a tested synthetic pathogen, with TSR performance quantified by the Relative Mean Absolute Bias (RMAB; x-axis, averaged across replicates) and the Relative Mean Standard Error (RMSE; y-axis, averaged across replicates). The color scale represents the average power, defined as the probability that the estimated parameter correctly recovers the sign of the true effect. The grey points indicate the control model’s performance. The parameters used to generate the fitted simulations were: *δ*_*Te*_ = *δ*_*RH*_ = −0.04, *ρ*_*M*_ = 50*%, σ*_*β*_ = 0*%, ρ*_*K*_ = 10*%*.

The estimates for relative humidity were more accurate (mean RMAB range: 15% in Rome to 31% in Rio de Janeiro) and precise (mean RMSE range: 11% in Rome to 20% in Rio de Janeiro), though this performance remains arguably poor given our simplified simulation and estimation protocol. The estimation of relative humidity effects had higher power, ranging from 93% in Rio de Janeiro to 100% in Dubai and Rome. There was also variability in accuracy across (*σ*_*I*_ range for the RMAB: 6–10%) and within (*σ*_*J*_ range for the RMAB: 10–17%) scenarios, although it was less pronounced than for temperature effects (Supplementary Table S5). For both weather variables, TSR performance varied across the simulated pathogens’ characteristics, though no consistent association was found (Figs. S3–S4). This higher performance compared to temperature likely reflects the greater variability of relative humidity and its effect on transmission across the four climates considered (Supplementary Table S1, Supplementary Figs. S5-S6).

The estimation performance for the other model versions is shown in Supplementary Figs. S7– S11 and Supplementary Tables S6–S9. Increasing the observation noise led to more inaccurate and imprecise estimates (with all values reported as mean ± SD) at each location (Supplementary Fig. S7), for both temperature (mean RMAB increase of 8% ± 5%, mean RMSE increase of 9% ± 3%) and relative humidity (mean RMAB increase of 6% ± 4%, mean RMSE increase of 3% ± 1%). Similarly, decreasing the effects of weather generally led to worse performance (Supplementary Fig. S8) (mean RMAB increase of 10% ± 25%, mean RMSE increase of 7% ± 16% for temperature; mean RMAB increase of 10% ± 12%, mean RMSE increase of 5% ± 3% for relative humidity). In contrast, adding process noise or term-time forcing did not, on average, worsen model performance (Supplementary Fig. S9) (mean RMAB change of 0% ± 9%, mean RMSE change of 0% ± 4% for temperature; mean RMAB decrease of 1% ± 4%, mean RMSE change of 0% ± 1% for relative humidity). However, greater variability across scenarios was observed with term-time forcing, with higher bias in some cases (Supplementary Fig. S10) (mean RMAB increase of 1% ± 20%, mean RMSE increase of 3% ± 13% for temperature; mean RMAB increase of 1% ± 10%, mean RMSE increase of 1% ± 4% for relative humidity).

Finally, in a sensitivity analysis, we found that changing the maximum dimension of the time smooth did not affect the estimates, with average differences in performance of 0% and 7% in RMAB and 1% and 0% for RMSE, respectively, for temperature and relative humidity across locations (Supplementary Fig. S11).

## 5 Discussion

The main goal of this study was to evaluate the reliability of TSR models in estimating the effects of weather on infectious diseases from population-level incidence data. Through a comprehensive simulation study based on a simple transmission model, we systematically assessed the accuracy and precision of TSR model formulations across four climates and fifty synthetic pathogens. Overall, we found that TSR generally performed poorly, with high bias and low precision in the effect estimates, especially for temperature. More broadly, our findings emphasize the importance of causal thinking and simulation-based experiments in evaluating study designs in infectious disease epidemiology.

Due to its simplicity and applicability, the TSR is highly popular in environmental epidemiology [26–28,30,31,34–40,42–54]. However, earlier research identified potential issues with this design, including challenges in modeling temporal changes in the immune population, autocorrelation due to transmission, and overdispersion [9]. In our study, these complexities were included through a causal framework represented by a simple SIRS model that captured transmission and susceptible depletion, augmented with an overdispersed (negative binomial) observation model. We deliberately used this simple model because it yielded a control TSR model that recovered the true weather effects in the absence of unobserved variables (Supplementary Fig.S4). In contrast, in realistic scenarios with unobserved variables affecting disease dynamics, TSR model estimates were substantially biased. This inaccuracy is striking, given our simplified modeling framework—in which a single regression coefficient represents the effect of each weather variable—and our use of advanced regression models that permit the estimation of flexible, nonlinear functions of time. Hence, our results caution against the indiscriminate use of TSR models for estimating the effects of weather on disease dynamics.

Importantly, the TSR test model’s performance may have been artificially inflated in our setting. This is because in our model *C*_*t*_ ≈ *I*_*t*_, due to the fixed generation time, an equality that does not hold in more realistic transmission models with distributed generation times. This consideration further reinforces our main point about the general unreliability of TSR models. More broadly, we propose that the choice of covariates in TSR models should be guided by causal models of the disease’s natural history and other data-generating mechanisms, rather than by more general considerations (e.g., autocorrelation or seasonality [9,10]).

Among all the control parameters tested, the level of measurement noise had the greatest impact on TSR model performance. Even low values—below the levels reported in the disease modeling literature—significantly reduced the accuracy and precision of the estimates. These findings align with earlier research that highlights measurement noise as a major issue in environmental epidemiology [9]. From a theoretical standpoint, we hypothesize this result is explained by high-frequency weather variability and the fact that, for directly transmitted infections (as modeled here), weather effects on transmission are immediate. As a result, the weather signature is characterized by high-frequency variations in the incidence data, and this signal may be lost by even modest levels of observation noise in the presence of unobserved variables.

Other influential control parameters included those describing the pathogen. Indeed, our analysis showed marked variability in TSR performance across the two main control parameters (mean age at infection and average duration of immunity) and the resulting basic reproduction number. This result was expected based on epidemiological theory, as these parameters critically influence transmission dynamics, particularly the amplitude, timing, and periodicity of epidemic peaks [55]. However, we were unable to find any clear link between these characteristics and TSR accuracy (Supplementary Figs. S7-S8). In practice, if these characteristics can be identified with reasonable confidence before analysis, we recommend conducting a simulation study similar to ours to assess, a priori, how TSR might perform for the system under investigation.

In contrast, other control parameters had a lower impact on TSR performance. In particular, adding process noise increased the bias only marginally in our application. This result was unexpected because, unlike measurement noise, process noise affects the transmission dynamics at every time point, causing its effects to accumulate and compound over time. Nevertheless, one would expect even stronger process and measurement noise in real-world applications.

We acknowledge several limitations of our study. First, we assumed that only two weather variables affected transmission. This choice was motivated by experimental evidence showing the independent effects of temperature and relative humidity on pathogen survival time, e.g., for influenza and SARS-CoV-2 [2,3]. Yet other causal variables—such as UV radiation [56]—may be biologically plausible, but because of the high correlations among weather variables, including them would likely make TSR even less reliable. Second, our intentionally simple transmission model overlooked several real-world complexities, such as the possibility that weather effects could be lagged (due to variability in the generation time among individuals) and non-monotonic (e.g., a U-shaped relationship with relative humidity, as indicated by some experimental evidence [2]). Extensions of GAMs have been developed to model lagged effects (e.g., distributed lag nonlinear models [57]), but we note that these were not needed here because we assumed a simple 1-week lag structure. More broadly, accounting for these additional complexities would make the estimation problem more difficult and likely reduce TSR performance even more.

Third, the outcomes considered here were time-series data on incident cases, but other outcomes can be defined and analyzed using TSR. This includes, in particular, the effective reproduction number, which can be reconstructed from the observed incidence rate and then regressed against weather variables of interest [42,47,56]. TSR performance might improve for this outcome, as the underlying causal diagram involves fewer unobserved variables [12]. However, this advantage could be offset by the smoothing introduced by the reconstruction. Although beyond the scope of this study, future research could investigate this question through a simulation study.

Finally, we considered only one TSR model; however, other formulations have been proposed to model the unobserved variables. For example, instead of the time-smooth covariate used here, Imai et al. suggested adding a covariate representing the count of past cases as a proxy for the immune population [9]. However, this approach would require detailed information on the duration of immunity, including both its average and its full distribution; therefore, its feasibility is uncertain if such information is unavailable. Future research could explore whether carefully constructed covariates can improve TSR performance. Still, we note that most TSR applications employed simple covariates similar to those in our analysis [10].

In conclusion, our findings indicate that, despite its popularity and ease of implementation, the TSR design can be unreliable, even with flexible regression-based statistical methods. These limitations highlight the complexity of infectious disease dynamics, including nonlinearities, various types of noise, and rapidly changing, yet unobserved, variables. Therefore, alternative methods—such as full transmission models or hybrid approaches, such as the TSIR model—may be necessary to clarify the effects of weather and, by extension, climate, on infectious diseases. More broadly, our results emphasize the value of simulation studies grounded in causal reasoning about the data-generating process for assessing existing methods under “experimental” conditions.

## Supporting information

Supplementary tables, text and images

## 7 Paper information

### Data and code availability statement

All code used to generate the simulations, perform the analysis, and produce the figures is currently stored in a private GitHub repository and will be made publicly available upon publication.

### Funding

This study was funded by the Max Planck Society.

### Contributions

P.G. and M.D.d.C. conceptualized the project and designed the methods. P.G. implemented the model and carried out the analyses. P.G. and M.D.d.C., with the support of C.K., L.A.B.G. and S.C.K., wrote the paper. M.D.d.C. supervised the project. All authors approved the final version of the paper.

## Acknowledgments

Computations were performed using the Max Planck Computing and Data Facility (MPCDF) high-performance computing cluster.

