## Supplementary tables, text and images for "The pitfalls of incidence-based time series regression for inferring the effects of weather on infectious diseases"

### Supplementary text

#### Variability across and within scenarios

Here, we derive the formulas for the variance across scenarios and the within-scenario variance across replicates reported in the main text.

Denote by $x_{ij}$ either RMAB or RMSE for scenario $i=1,\cdots,n_{I}$ and within-scenario replicate $j=1,\cdots,n_{J}$.

The mean for scenario $i$ is defined as

$\bar{x}_{i}=\frac{1}{n_{J}}\sum_{j=1}^{n_{J}} x_{ij}$,

the grand mean across scenarios is

$\bar{x}=\frac{1}{n_{I}}\sum_{i=1}^{n_{I}} \bar{x}_{i}$.

The overall variance of $x_{ij}$

$$\sigma^{2}=V(x_{ij})$$

can be decomposed as:

$\sigma^{2}=\sigma_{I}^{2}+\sigma_{J}^{2}$.

The first term represents the variance across scenarios

$\sigma_{I}^{2}=E[(\bar{x}-\bar{x}_{i})^{2}]$,

whereas the second represents variance across replicates within scenarios

$$\sigma_{J}^{2}=E[(x_{ij}-\bar{x}_{i})^{2}]$$

### Supplementary figures

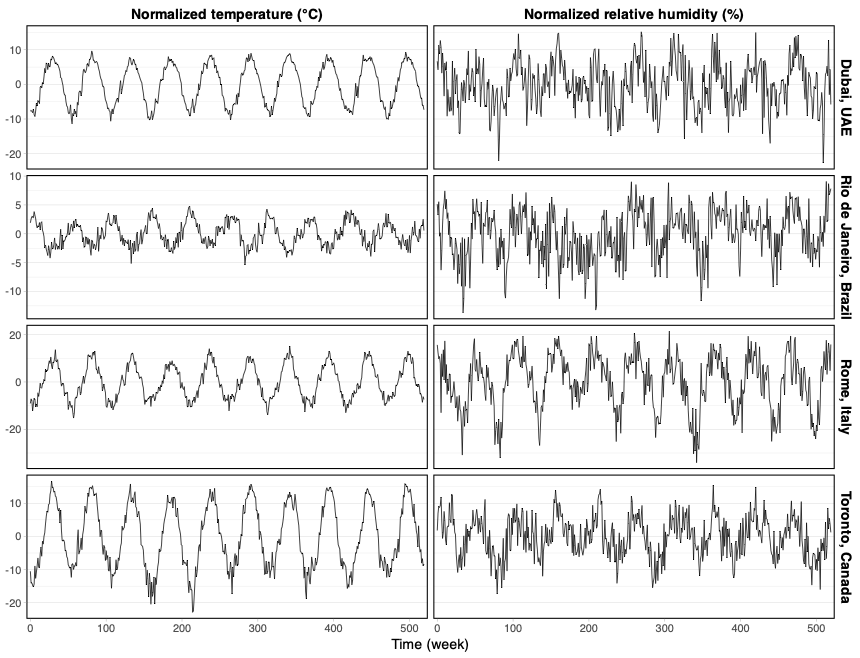
**Figure S1: Time plot of weather variables.** The normalized temperature is defined as $Te^{'}=Te_{t}-\bar{Te}$ , and the normalized relative humidity as $RH^{'}=RH_{t}- \bar{RH}$.

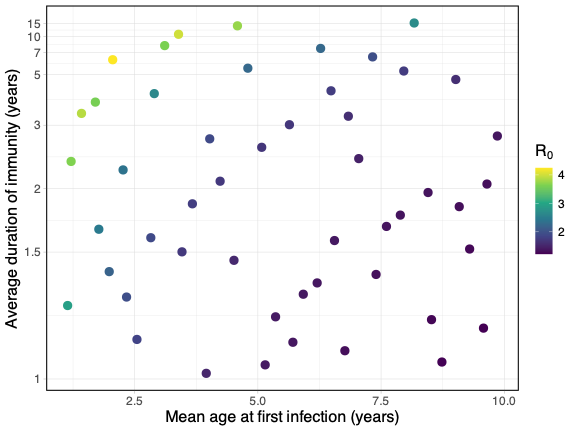
**Figure S2: Characteristics of the synthetic pathogens.** Combination of sampled values of mean age at first infection ($A$, x-axis) and average duration of immunity ($\alpha$, y-axis). The filling color corresponds to the respective basic reproduction number, which can be obtained from the combination of the two pathogen-specific parameters $R_{0}=1+\frac{1+\left( \alpha+ \mu\right)^{-1}}{A\left( \mu+ 1 \right)}$.

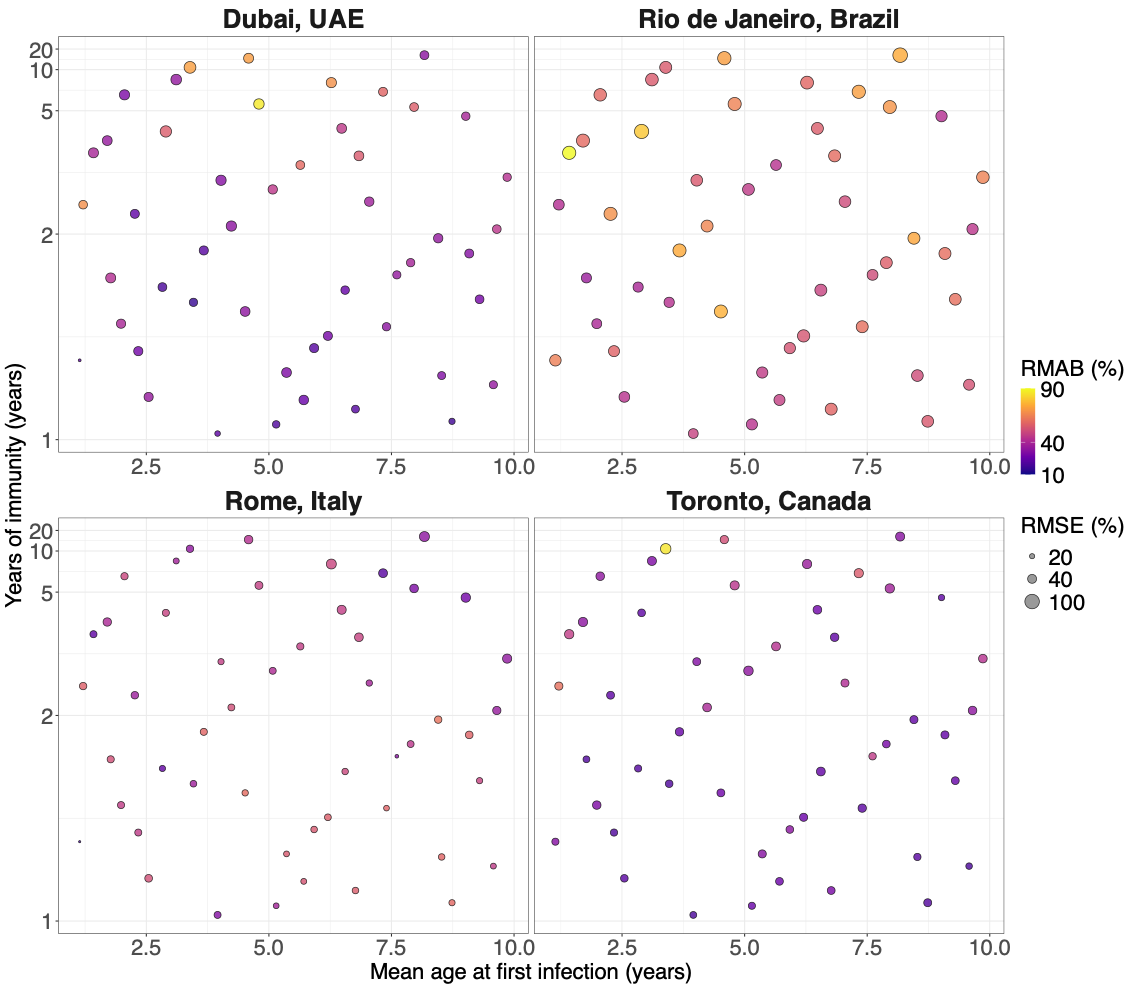
**Figure S3: Temperature performance distribution across scenarios for the test model.** The color scale shows the Relative Mean Absolute Bias (RMAB) of temperature effect estimates across the replicates of each scenario. The size of the points indicates the Relative Mean Standard Error (RMSE) for each scenario.

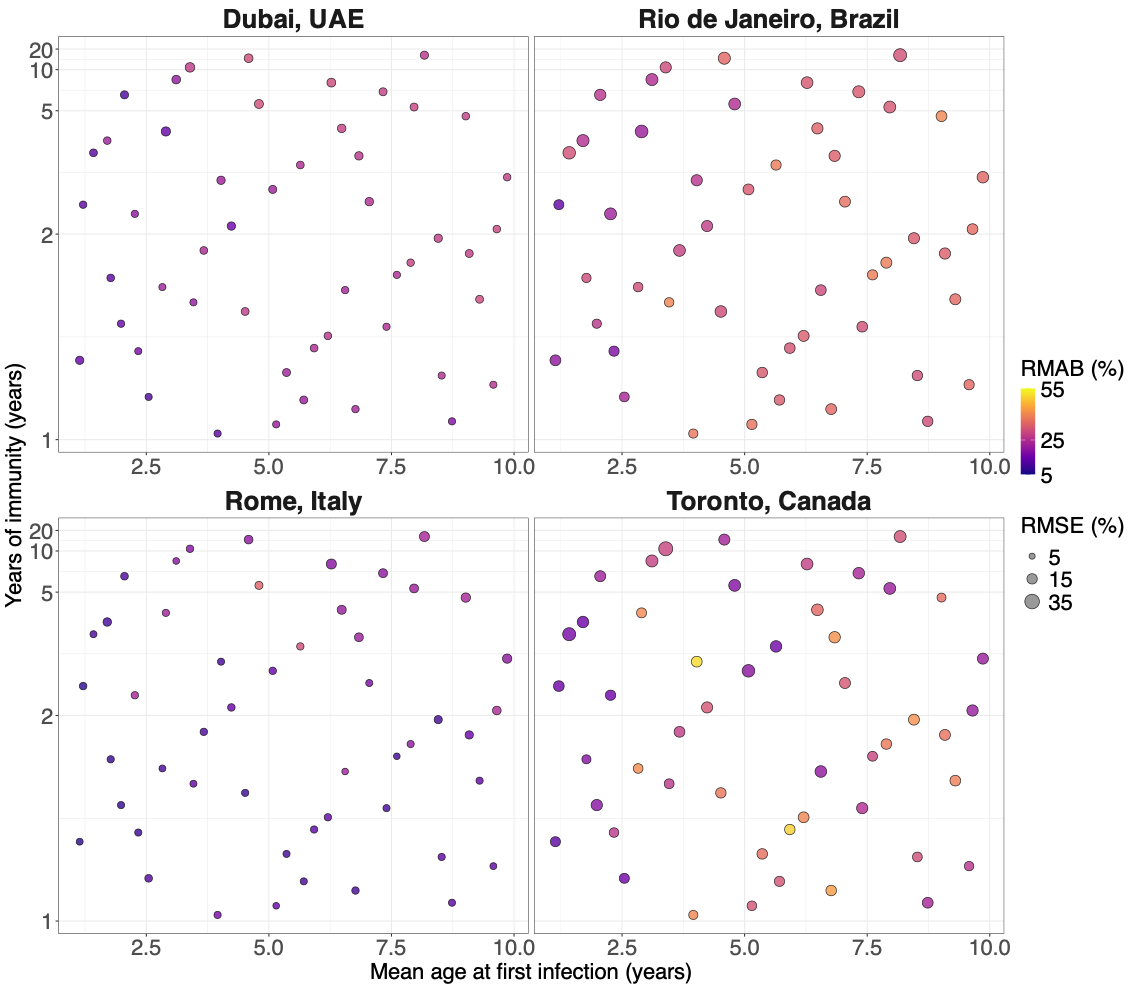
**Figure S4: Relative humidity performance distribution across scenarios for the test model.** The color scale shows the Relative Mean Absolute Bias (RMAB) of relative humidity effect estimates across the replicates of each scenario. The size of the points indicates the Relative Mean Standard Error (RMSE) for each scenario.

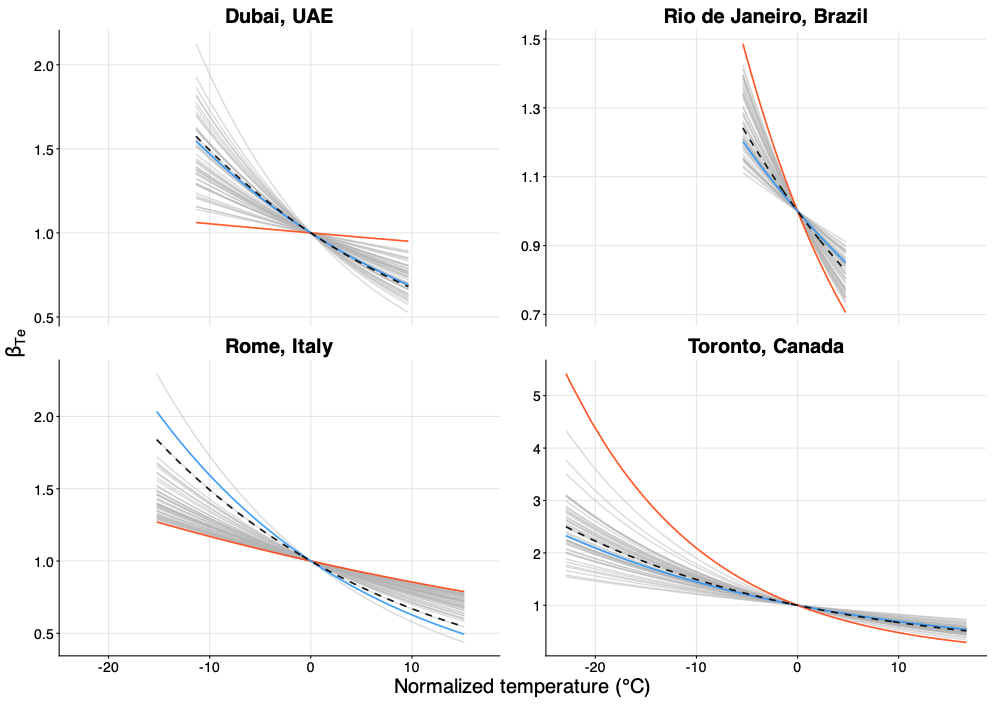
**Figure S5: Temperature component of the seasonal transmissibility rate.** The grey lines show the component of the transmission coefficient due to temperature, obtained using scenario-averaged effect estimates (on the y-axis), as a function of the normalized temperature (on the x-axis). Averages with the lowest and highest RMAB in each location are indicated in blue and red, respectively. Dashed lines represent the ground-truth relationship used in the generation process.

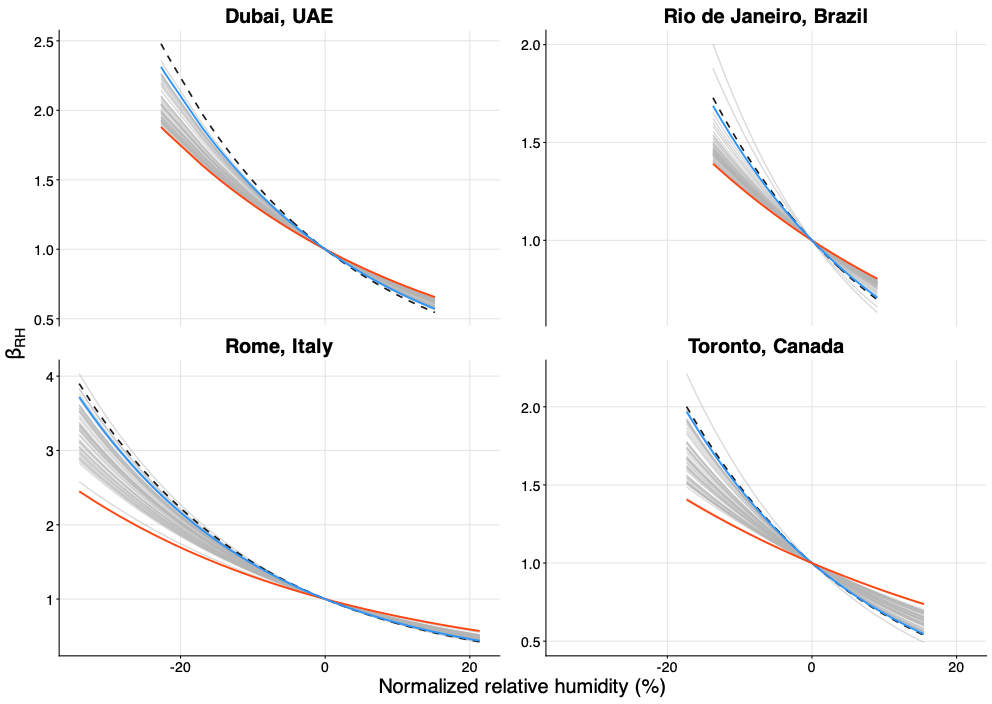
**Figure S6: Relative humidity component of the seasonal transmissibility rate.** The grey lines show the component of the transmission coefficient due to relative humidity, obtained using scenario-averaged effect estimates (on the y-axis), as a function of the normalized relative humidity (on the x-axis). The averages with the lowest and highest RMAB in each location are indicated in blue and red, respectively. Dashed lines represent the ground-truth relationship used in the generation process

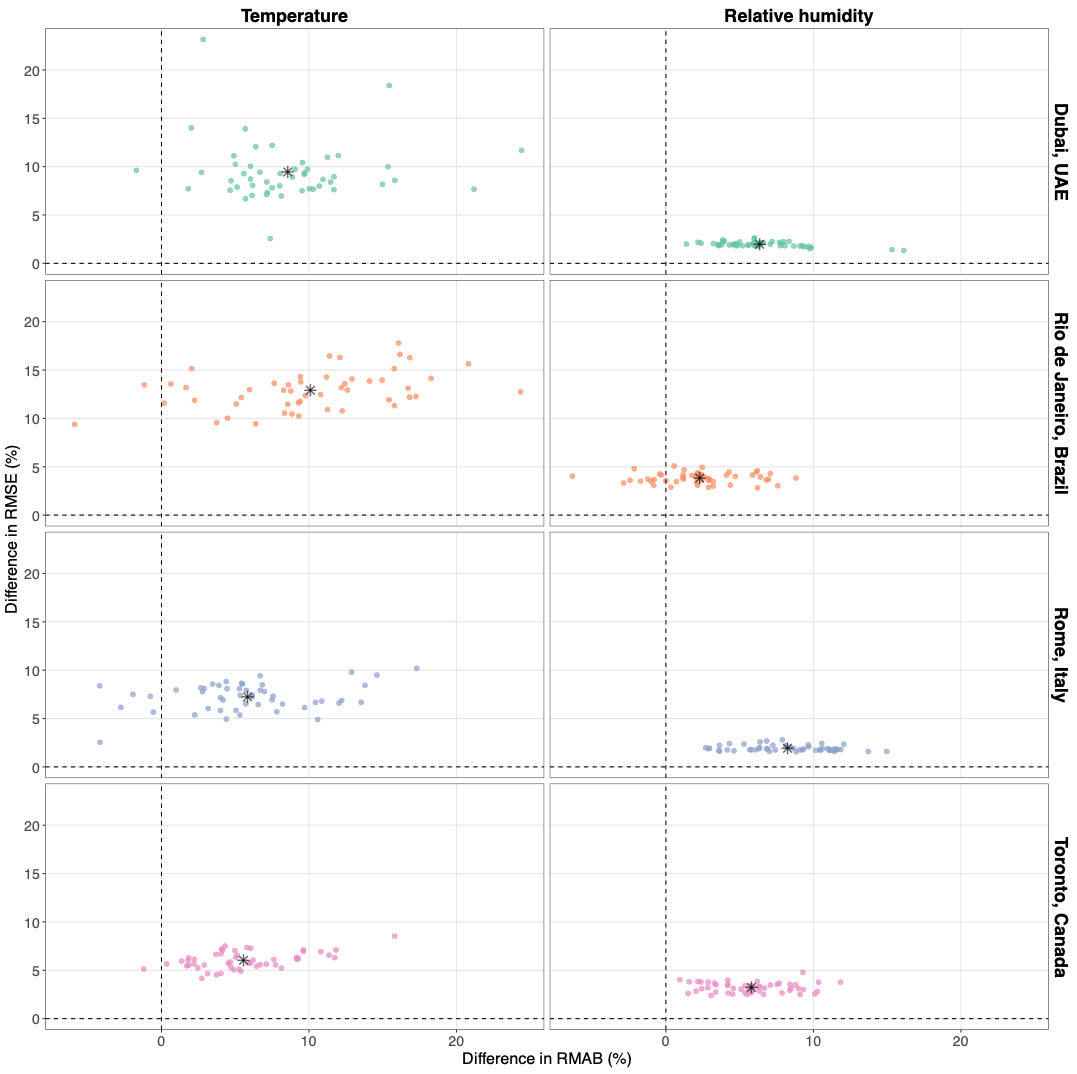
**Figure S7: Difference in performance when increasing measurement noise.** Across different locations, each point represents one of the 50 scenarios, with the scenario-coupled differences in Relative Mean Absolute Bias (RMAB) on the x-axis and Relative Mean Standard Error (RMSE) on the y-axis at varying measurement noise levels. The black asterisk denotes the grand mean of RMAB and RMSE across scenarios. The differences are always taken as “measurement noise = 14% (treatment) − measurement noise = 10% (baseline)”; a positive difference indicates a worsening of the performance measure with the change of the analyzed parameter.

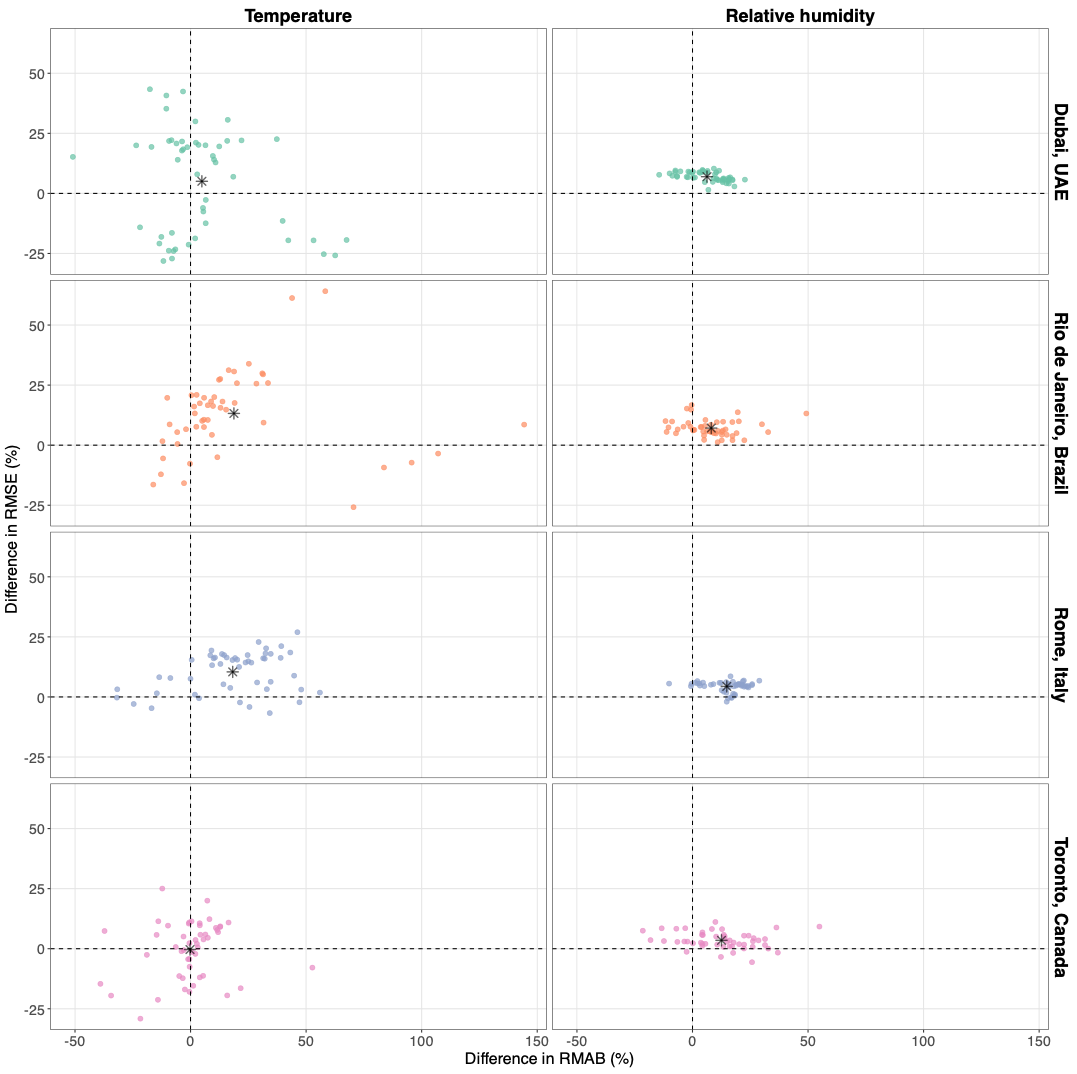
**Figure S8: Difference in performance when decreasing climate effects.** Across different locations, each point represents one of the 50 scenarios, with the scenario-coupled differences in Relative Mean Absolute Bias on the x-axis and Relative Mean Standard Error on the y-axis at varying levels of climate effect. The black asterisk denotes the grand mean of RMAB and RMSE across scenarios. The differences are taken as “δ = −0.02 (treatment) − δ = −0.04 (baseline)”; a positive difference indicates a worsening of the performance measure with the change of the analyzed parameter.

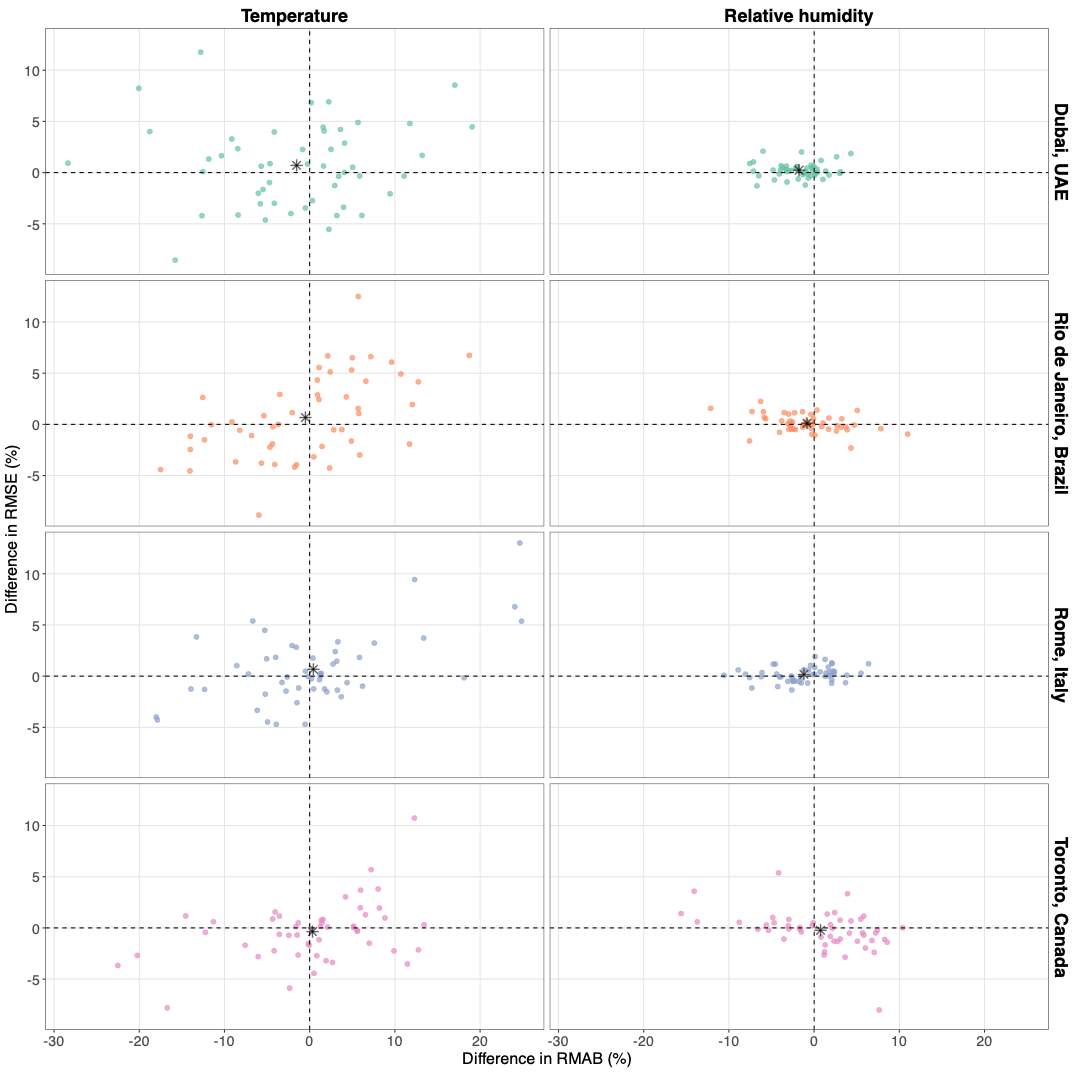

**Figure S9: Difference in performance when increasing process noise.** Across different locations, each point represents one of the 50 scenarios, with the scenario-coupled differences in Relative Mean Absolute Bias on the x-axis and Relative Mean Standard Error on the y-axis at varying process noise levels. The black asterisk denotes the grand mean of RMAB and RMSE across scenarios. The differences are always taken as “process noise = 5% (treatment) − process noise = 0% (baseline)”; a positive difference indicates a worsening of the performance measure with the change of the analyzed parameter.

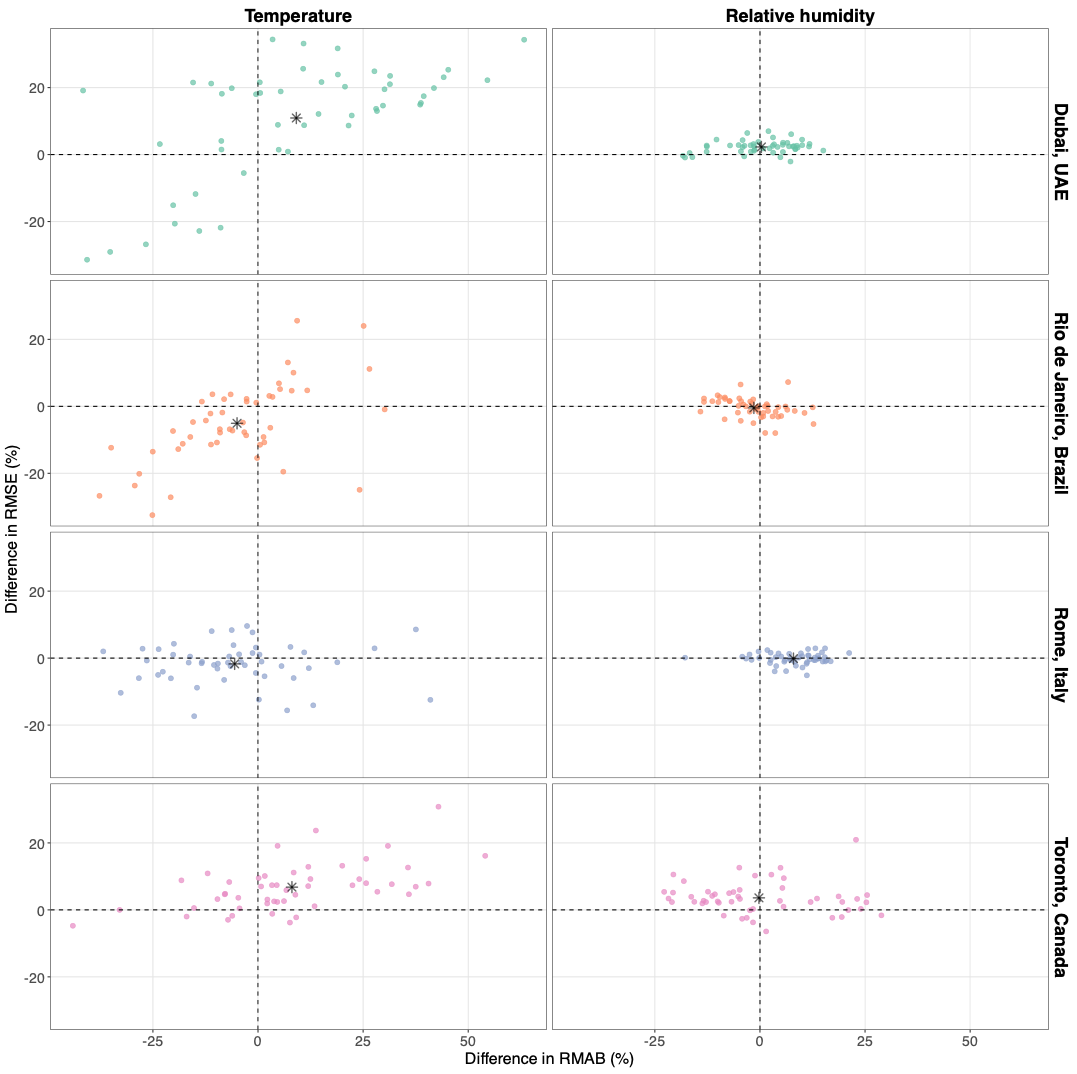
**Figure S10: Difference in performance when including term-time forcing.** Across different locations, each point represents one of the 50 scenarios, with the scenario-coupled differences in Relative Mean Absolute Bias on the x-axis and Relative Mean Standard Error on the y-axis, with and without term-time forcing. The black asterisk denotes the grand mean of RMAB and RMSE across scenarios. The differences are taken as “with term-time forcing (treatment) − without term-time forcing (baseline)”; a positive difference indicates a worsening of the performance measure with the change of the analyzed parameter.

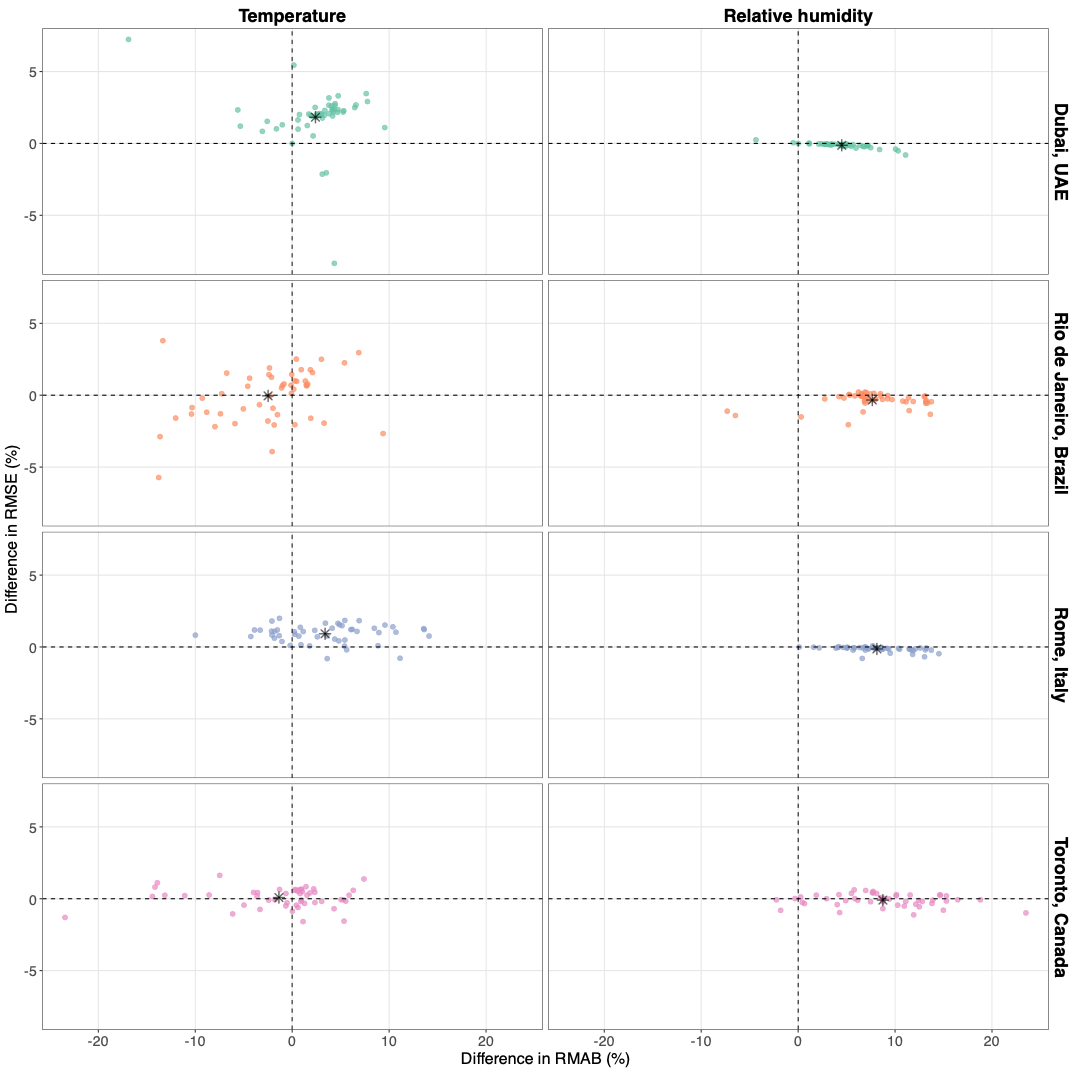
**Figure S11: Difference in performance when increasing the smoother dimension.** Across different locations, each point represents one of the 50 scenarios, with the scenario-coupled differences in Relative Mean Absolute Bias on the x-axis and Relative Mean Standard Error on the y-axis, with and without term-time forcing. The black asterisk denotes the grand mean of RMAB and RMSE across scenarios. The difference is taken as “the performance measure using a maximum of 75 degrees of freedom (treatment) minus the one obtained using 52 degrees of freedom (baseline)” for the GAM spline smooth; a positive difference indicates a worsening of the performance measure with the change of the analyzed parameter.

### Supplementary tables

| **Location** | **Köppen-Geiger code** | **Brief description** | **Temperature (°C) standard deviation** | **Relative humidity (%)**  **standard deviation** |
| --- | --- | --- | --- | --- |
| Rio de Janeiro, Brazil | Aw | Tropical wet and dry climate | 2.0 | 4.5 |
| Dubai, United Arab Emirates | BWh | Hot desert climate | 5.7 | 6.9 |
| Rome, Italy | Csa | Hot-summer Mediterranean climate | 7.2 | 11.4 |
| Toronto, Canada | Dfb | Humid continental climate | 9.3 | 6.5 |

**Table S1: Climate characteristics in the four selected locations.**

| **Parameter** | **Meaning** | **Value** |
| --- | --- | --- |
| $\mu$ | Birth/death rate (individuals/week) | $\frac{1}{80\cdot52}$ |
| $N$ | Fixed population size (individuals) | $5\cdot10^{6}$ |
| $\alpha^{-1}$ | Average duration of protection (year) | 1 to 20 |
| A | Mean age at first infection (year) | 1 to 10 |
| $R_{0}$ | Basic reproduction number | 1.22 to 4.25 |
| $\gamma^{-1}$ | Generation time (week) | 1 |
| $\delta_{Te}$ | Effect of temperature on transmission | $-0.02, -0.04$ |
| $\delta_{RH}$ | Effect of relative humidity on transmission | $-0.02, -0.04$ |
| $\rho_{M}$ | Average reporting probability | $0.5$ |
| $\rho_{k}$ | Measurement noise | $0.1$, 0.16 |
| $\sigma_{\beta}$ | Process noise | $0,0.05$ |
| $\delta_{school}$ | Effect of Term-time on transmissibility | 0.18 |
| $\iota$ | Absolute number of imported infected individuals | ${10}^{-5}$ |

[**Table S**](https://docs.google.com/document/d/13k2twcDeKcA-crf_TAYJUgoqn5jhSEb9L437dt5NVWs/edit#table_parameters)**2: Transmission model parameters.**

| **Study** | **Pathogen** | **Location, Period** | **Observation frequency** | **Estimate of** $\rho_{\boldsymbol{k}}$ | **Source** |
| --- | --- | --- | --- | --- | --- |
| Lau et al., Epidemics 2024 [1] | RSV | Hong Kong, 2014–2019 | Weekly | 0.33 | Table S2 |
|  | RSV | Hong Kong, 2020–2024 | Weekly | 1.28 |  |
|  | RSV | South Korea, 2014–2019 | Weekly | 0.44 |  |
|  | RSV | South Korea, 2020–2024 | Weekly | 0.69 |  |
|  | HFMD | Hong Kong, 2014–2019 | Weekly | 0.62 |  |
|  | HFMD | Hong Kong, 2020–2024 | Weekly | 0.49 |  |
|  | HFMD | South Korea, 2014–2019 | Weekly | 0.27 |  |
|  | HFMD | South Korea, 2020–2024 | Weekly | 0.32 |  |
| Becker et al., PNAS 2016 [2] | Measles | London, 1904 | Weekly | 0.85, 0.92 | Main text |
| Baracchini et al., Advances in Water Resources 2017 [3] | Cholera | Dhaka, Patna, Midnapore,  1892-1941 | Weekly | 0.82, 0.79, 0.52 | Main text |
| Laneri et al., Plos Comp. Bio 2010 [4] | Malaria | Kutch, India (1987 - 2007);  Barmer, India (1985 - 2005) | Monthly | 0.613, 0.598 | Table S4 |
| Shah et al., R Soc Open Sci. 2022 [5] | Mumps | Harvard (February-September 2016) | Weekly | 0.54 (0.36,0.72) | Table 1 |

**Table S3: Mini-review of previously estimated measurement noise.** The table above summarizes some previously estimated measurement noise parameters found in the literature.

| **Location** | **Metric** | **Temperature** | **Relative humidity** |
| --- | --- | --- | --- |
| Dubai, UAE | RMAB | 8 (1) [6] | 6 (1) [5] |
|  | RMSE | 10 (1) [0] | 8 (1) [0] |
| Rio de Janeiro, Brazil | RMAB | 23 (4) [18] | 10 (2) [8] |
|  | RMSE | 28 (5) [1] | 13 (2) [1] |
| Rome, Italy | RMAB | 8 (2) [6] | 5 (1) [4] |
|  | RMSE | 11 (2) [1] | 7 (1) [0] |
| Toronto, Canada | RMAB | 7 (1) [5] | 10 (2) [8] |
|  | RMSE | 8 (1) [0] | 13 (2) [1] |

**Table S4 Mean RMAB and MSE for the control model with the baseline parameter set.** The table above shows the performances for all locations with $\rho_{k}=10\%, \delta_{Te}=\delta_{RH}=-0.04, \sigma_{\beta}=0\%, \rho_{M}=50\%$. Each cell corresponds to one variable and one location and displays, first, the RMAB (%) across all replicates and scenarios, then its inter-scenario variability ($\sigma_{I}$) in round brackets, and finally its intra-scenario variability ($\sigma_{J}$) in square brackets. Below, the respective RMSE (%) across all replicates and scenarios is shown, along with its inter- and intra-scenario variability.

| **Location** | **Metric** | **Temperature** | **Relative humidity** |
| --- | --- | --- | --- |
| Dubai, UAE | RMAB | 38 (16) [24] | 22 (6) [10] |
|  | RMSE | 42 (9) [5] | 11 (1) [0] |
| Rio de Janeiro, Brazil | RMAB | 57 (12) [42] | 31 (6) [17] |
|  | RMSE | 68 (12) [4] | 20 (3) [1] |
| Rome, Italy | RMAB | 44 (11) [22] | 15 (6) [10] |
|  | RMSE | 29 (7) [3] | 11 (2) [1] |
| Toronto, Canada | RMAB | 32 (13) [22] | 29 (10) [15] |
|  | RMSE | 35 (6) [2] | 20 (4) [1] |

**Table S5 Mean RMAB and RMSE for the tested model with the baseline parameter set.** The table above shows the performances for all locations, with $\rho_{k}=10\%, \delta_{Te}=\delta_{RH}=-0.04, \sigma_{\beta}=0\%,\rho_{M}=50\%$. Each cell corresponds to one variable and one location and displays, first, the RMAB (%) across all replicates and scenarios, then its inter-scenario variability ($\sigma_{I}$) in round brackets, and finally its intra-scenario variability ($\sigma_{J}$) in square brackets. Below, the respective RMSE (%) across all replicates and scenarios is shown, along with its inter- and intra-scenario variability.

| **Location** | **Metric** | **Temperature** | **Relative humidity** |
| --- | --- | --- | --- |
| Dubai, UAE | RMAB | 46 (18) [30] | 29 (6) [12] |
|  | RMSE | 51 (9) [4] | 13 (2) [1] |
| Rio de Janeiro, Brazil | RMAB | 67 (15) [50] | 33 (5) [20] |
|  | RMSE | 81 (13) [4] | 24 (4) [1] |
| Rome, Italy | RMAB | 50 (13) [27] | 23 (6) [12] |
|  | RMSE | 37 (7) [2] | 13 (2) [1] |
| Toronto, Canada | RMAB | 37 (14) [27] | 34 (11) [19] |
|  | RMSE | 41 (6) [2] | 23 (4) [1] |

**Table S6 Mean RMAB and RMSE for the tested model with increased measurement noise.** The table above shows the performances for all locations, with $\rho_{k}=16\%, \delta_{Te}=\delta_{RH}=-0.04, \sigma_{\beta}=0\%, \bar{\rho}=50\%$. Each cell corresponds to one variable and one location and displays, first, the RMAB (%) across all replicates and scenarios, then its inter-scenario variability ($\sigma_{I}$) in round brackets, and finally its intra-scenario variability ($\sigma_{J}$) in square brackets. Below, the respective RMSE (%) across all replicates and scenarios is shown, along with its inter- and intra-scenario variability.

| **Location** | **Metric** | **Temperature** | **Relative humidity** |
| --- | --- | --- | --- |
| Dubai, UAE | RMAB | 43 (24) [26] | 29 (12) [14] |
|  | RMSE | 47 (21) [8] | 18 (2) [1] |
| Rio de Janeiro, Brazil | RMAB | 75 (31) [49] | 39 (8) [23] |
|  | RMSE | 82 (19) [7] | 27 (3) [1] |
| Rome, Italy | RMAB | 62 (20) [33] | 30 (8) [13] |
|  | RMSE | 40 (6) [4] | 15 (1) [1] |
| Toronto, Canada | RMAB | 31 (11) [22] | 41 (13) [21] |
|  | RMSE | 35 (13) [5] | 23 (2) [1] |

**Table S7 Mean RMAB and RMSE for the tested model with decreased effect of climate.** The table above shows the performances for all locations, with $\rho_{k}=10\%, \delta_{Te}=\delta_{RH}=-0.02, \sigma_{\beta}=0\%, \bar{\rho}=50\%$. Each cell corresponds to one variable and one location and displays, first, the RMAB (%) across all replicates and scenarios, then its inter-scenario variability ($\sigma_{I}$) in round brackets, and finally its intra-scenario variability ($\sigma_{J}$) in square brackets. Below, the respective RMSE (%) across all replicates and scenarios is shown, along with its inter- and intra-scenario variability.

| **Location** | **Metric** | **Temperature** | **Relative humidity** |
| --- | --- | --- | --- |
| Dubai, UAE | RMAB | 36 (10) [27] | 21 (5) [11] |
|  | RMSE | 42 (9) [9] | 11 (1) [1] |
| Rio de Janeiro, Brazil | RMAB | 56 (9) [43] | 30 (5) [18] |
|  | RMSE | 69 (10) [7] | 20 (3) [2] |
| Rome, Italy | RMAB | 44 (7) [24] | 14 (4) [10] |
|  | RMSE | 30 (6) [5] | 11 (2) [1] |
| Toronto, Canada | RMAB | 32 (8) [24] | 29 (6) [17] |
|  | RMSE | 35 (4) [4] | 20 (3) [2] |

**Table S8 Mean RMAB and RMSE for the tested model with increased process noise.** The table above shows the performances for all locations, with $\rho_{k}=10\%, \delta_{Te}=\delta_{RH}=-0.04, \sigma_{\beta}=5\%, \bar{\rho}=50\%$. Each cell corresponds to one variable and one location and displays, first, the RMAB (%) across all replicates and scenarios, then its inter-scenario variability ($\sigma_{I}$) in round brackets, and finally its intra-scenario variability ($\sigma_{J}$) in square brackets. Below, the respective RMSE (%) across all replicates and scenarios is shown, along with its inter- and intra-scenario variability.

| **Location** | **Metric** | **Temperature** | **Relative humidity** |
| --- | --- | --- | --- |
| Dubai, UAE | RMAB | 47 (21) [35] | 23 (8) [12] |
|  | RMSE | 53 (18) [7] | 13 (2) [1] |
| Rio de Janeiro, Brazil | RMAB | 52 (16) [39] | 29 (7) [16] |
|  | RMSE | 63 (14) [3] | 20 (4) [1] |
| Rome, Italy | RMAB | 38 (13) [20] | 23 (6) [11] |
|  | RMSE | 28 (5) [2] | 11 (2) [0] |
| Toronto, Canada | RMAB | 40 (14) [27] | 28 (11) [18] |
|  | RMSE | 42 (7) [2] | 23 (5) [1] |

**Table S9 Mean RMAB and RMSE for the tested model, including Term-time forcing.** The table above shows the performances for all locations, with $\rho_{k}=10\%, \delta_{Te}=\delta_{RH}=-0.04, \sigma_{\beta}=0\%, \bar{\rho}=50\%$. Each cell corresponds to one variable and one location and displays, first, the RMAB (%) across all replicates and scenarios, then its inter-scenario variability ($\sigma_{I}$) in round brackets, and finally its intra-scenario variability ($\sigma_{J}$) in square brackets. Below, the respective RMSE (%) across all replicates and scenarios is shown, along with its inter- and intra-scenario variability.
